# Effect of prior infection, vaccination, and hybrid immunity against symptomatic BA.1 and BA.2 Omicron infections and severe COVID-19 in Qatar

**DOI:** 10.1101/2022.03.22.22272745

**Authors:** Heba N. Altarawneh, Hiam Chemaitelly, Houssein H. Ayoub, Patrick Tang, Mohammad R. Hasan, Hadi M. Yassine, Hebah A. Al-Khatib, Maria K. Smatti, Peter Coyle, Zaina Al-Kanaani, Einas Al-Kuwari, Andrew Jeremijenko, Anvar Hassan Kaleeckal, Ali Nizar Latif, Riyazuddin Mohammad Shaik, Hanan F. Abdul-Rahim, Gheyath K. Nasrallah, Mohamed Ghaith Al-Kuwari, Adeel A. Butt, Hamad Eid Al-Romaihi, Mohamed H. Al-Thani, Abdullatif Al-Khal, Roberto Bertollini, Laith J. Abu-Raddad

## Abstract

**BACKGROUND:** Protection offered by five different forms of immunity, combining natural and vaccine immunity, was investigated against SARS-CoV-2 Omicron symptomatic BA.1 infection, symptomatic BA.2 infection, BA.1 hospitalization and death, and BA.2 hospitalization and death, in Qatar, between December 23, 2021 and February 21, 2022.

**METHODS:** Six national, matched, test-negative case-control studies were conducted to estimate effectiveness of BNT162b2 (Pfizer-BioNTech) vaccine, mRNA-1273 (Moderna) vaccine, natural immunity due to prior infection with pre-Omicron variants, and hybrid immunity from prior infection and vaccination.

**RESULTS:** Effectiveness of only prior infection against symptomatic BA.2 infection was 46.1% (95% CI: 39.5-51.9%). Effectiveness of only two-dose BNT162b2 vaccination was negligible at -1.1% (95% CI: -7.1-4.6), but nearly all individuals had received their second dose several months earlier. Effectiveness of only three-dose BNT162b2 vaccination was 52.2% (95% CI: 48.1-55.9%). Effectiveness of hybrid immunity of prior infection and two-dose BNT162b2 vaccination was 55.1% (95% CI: 50.9-58.9%). Effectiveness of hybrid immunity of prior infection and three-dose BNT162b2 vaccination was 77.3% (95% CI: 72.4-81.4%). Meanwhile, prior infection, BNT162b2 vaccination, and hybrid immunity all showed strong effectiveness >70% against any severe, critical, or fatal COVID-19 due to BA.2 infection. Similar levels and patterns of effectiveness were observed for BA.1 and for the mRNA-1273 vaccine.

**CONCLUSIONS:** There are no discernable differences in the effects of prior infection, vaccination, and hybrid immunity against BA.1 versus BA.2. Hybrid immunity resulting from prior infection and recent booster vaccination confers the strongest protection against either subvariant. Vaccination enhances protection of those with a prior infection.

## Introduction

Qatar endured a severe acute respiratory syndrome coronavirus 2 (SARS-CoV-2) Omicron (B.1.1.529)^1^ wave that started on December 19, 2021 and peaked in mid-January, 2022.^2-6^ The wave was first dominated by the BA.1 Omicron subvariant, but within a few days, the BA.2 subvariant predominated (Figure S1 of Supplementary Appendix). While BA.1 and BA.2 remain classified as subvariants of the Omicron variant, there is considerable genetic distance between them.^7^ The protection of prior immunity against these subvariants, whether immunity is induced by prior infection, vaccination, or a hybrid of both, remains to be established.

Accordingly, we investigated the protection of prior infection with pre-Omicron variants: protection of the BNT162b2 (Pfizer-BioNTech)^8^ and mRNA-1273 (Moderna)^9^ mRNA coronavirus disease 2019 (COVID-19) vaccines, after the second dose and after the third/booster dose, and protection of hybrid immunity of prior infection and vaccination. Five different types of immunity protection were investigated against symptomatic BA.1 infection, symptomatic BA.2 infection, and any symptomatic Omicron infection with either subvariant, between December 23, 2021 and February 21, 2022. Protection was also investigated against any severe (acute-care hospitalization),^10^ critical (intensive-care-unit hospitalization),^10^ or fatal^11^ COVID-19 due to BA.1, BA.2, or any Omicron infection.

## Methods

### Study population and data sources

This study was conducted in the resident population of Qatar. It analyzed the national, federated databases for COVID-19 vaccination, laboratory testing, hospitalization, and death, retrieved from the integrated nationwide digital-health information platform. Databases include all SARS-CoV-2-related data and associated demographic information, with no missing information, since pandemic onset, documenting all polymerase chain reaction (PCR) tests and more recently, rapid antigen tests conducted at healthcare facilities (from January 5, 2022 onward).

Every PCR test (but not rapid antigen tests) conducted in Qatar is classified on the basis of symptoms and the reason for testing (clinical symptoms, contact tracing, surveys or random testing campaigns, individual requests, routine healthcare testing, pre-travel, at port of entry, or other). Qatar has unusually young, diverse demographics, in that only 9% of its residents are ≥50 years of age, and 89% are expatriates from over 150 countries.^12^ Qatar launched its COVID-19 vaccination program at the end of December of 2020 using both the BNT162b2 and mRNA-1273 mRNA vaccines.^13^ Nearly all individuals were vaccinated in Qatar, but if vaccinated elsewhere, those vaccinations were still recorded in the health system at the port of entry upon arrival in Qatar. Further descriptions of the study population and these national databases have been reported previously.^6,12-19^

### Study design

Two studies were conducted. In the first study, labeled as the BNT162b2-vaccine study, we estimated effectiveness of prior infection, BNT162b2 vaccination, and hybrid immunity of prior infection and BNT162b2 vaccination together against symptomatic BA.1 infection, symptomatic BA.2 infection, and any symptomatic Omicron infection due to either subvariant, using the test-negative, case-control study design, a standard design for assessing effectiveness of vaccination and prior infection.^3,18,20-23^ Effectiveness was also estimated against any severe,^10^ critical,^10^ or fatal^11^ COVID-19. In the second study, labeled as the mRNA-1273-vaccine study, the same effectiveness measures were estimated, but with mRNA-1273 vaccination instead of BNT162b2 vaccination.

For estimation of effectiveness against symptomatic infection, we exact-matched cases (PCR-positive persons) and controls (PCR-negative persons) identified between December 23, 2021 and February 21, 2022, during the Omicron wave,^2-6^ in a one-to-one ratio by sex, 10-year age group, nationality, and calendar week of PCR testing. Matching was done to control for known differences in the risk of exposure to SARS-CoV-2 infection in Qatar.^12,24-27^ Matching by these factors was previously shown to provide adequate control of differences in the risk of infection exposure in Qatar, in studies of different epidemiologic designs and that included control groups, including test-negative case-control studies.^13,14,18,28,29^ For estimation of effectiveness against any severe,^10^ critical,^10^ or fatal^11^ COVID-19, we used a matching ratio of one-to-five to improve statistical precision of estimates.

Only the first PCR-positive test during the study was included for each case, while all PCR-negative tests during the study were included for each control. Controls included individuals with no record of a PCR-positive test during the study period. Only PCR tests conducted for clinical suspicion among cases and controls, that is, due to symptoms compatible with a respiratory tract infection (symptomatic infection), were included in this analysis.

SARS-CoV-2 reinfection is conventionally defined as a documented infection ≥90 days after an earlier infection, to avoid misclassification of prolonged PCR positivity as reinfection, if a shorter time interval is used.^3,30,31^ Prior infection was thus defined as a PCR-positive test ≥90 days before this study’s PCR test. Individuals with a PCR-positive test <90 days before the study’s PCR test were excluded from both cases and controls. Accordingly, prior infections in this study were due to pre-Omicron variants, as they occurred prior to onset of the Omicron wave in Qatar.^2-6^

Individuals who did not receive the BNT162b2 or mRNA-1273 vaccines and those who received mixed vaccines were excluded from this analysis. Individuals who did not complete 14 days after the second vaccine dose, or did not complete 7 days after the third dose, were also excluded. These inclusion and exclusion criteria were implemented to allow adequate time for build-up of immunity after the second and booster doses,^6,17^ and to minimize different types of potential bias, as informed by earlier analyses in the same population.^14,29^ Every control that met the inclusion criteria and that could be matched to a case was included in analysis. PCR-test outcome, prior-infection status, and vaccination status were ascertained at the time of the PCR test.

Classification of COVID-19 case severity,^10^ criticality,^10^ and fatality^11^ followed World Health Organization guidelines, and assessments were made by trained medical personnel independent of the study investigators and using individual chart reviews, as part of a national protocol applied to every hospitalized COVID-19 patient. Details of COVID-19 severity, criticality, and fatality classifications are found in Section S1.

Every hospitalized COVID-19 patient underwent infection severity assessment every three days until discharge or death. We classified individuals who progressed to severe, critical, or fatal COVID-19 between the time of the PCR-positive test and the end of the study based on their worst disease outcome, starting with death,^11^ followed by critical disease,^10^ and then severe disease.^10^

### Laboratory methods and subvariant ascertainment

The large Omicron-wave exponential-growth phase started on December 19, 2021 and peaked in mid-January, 2022.^2-6^ A total of 315 random SARS-CoV-2-positive specimens collected between December 19, 2021 and January 22, 2022 were viral whole-genome sequenced on a Nanopore GridION sequencing device. Of these, 300 (95.2%) were confirmed as Omicron infections and 15 (4.8%) as Delta (B.1.617.2)^1^ infections.^2-6^ Of 286 Omicron infections with confirmed subvariant status, 68 (23.8%) were BA.1 cases and 218 (76.2%) were BA.2 cases.

Informed by viral genome sequencing and real-time reverse-transcription PCR (RT-qPCR) genotyping (further data are presented in Section S2), a SARS-CoV-2 infection with the Omicron BA.1 subvariant was proxied as an S-gene “target failure” (SGTF) case using the TaqPath COVID-19 Combo Kit (Thermo Fisher Scientific, USA) that tests for the S-gene and is affected by the del-69/70 mutation in the S-gene.^32^ A SARS-CoV-2 infection with the BA.2 subvariant was proxied as a non-SGTF case, using this TaqPath Kit.

Further details of laboratory methods for the RT-qPCR testing are found in Section S2. All PCR testing was conducted at the Hamad Medical Corporation Central Laboratory or at Sidra Medicine Laboratory, following standardized protocols.

### Statistical analysis

While all records of PCR testing in Qatar were examined for selection of cases and controls, only matched samples were analyzed. Cases and controls were described using frequency distributions and measures of central tendency, and compared using standardized mean differences (SMDs). An SMD was defined as the difference in the mean of a covariate between groups, divided by the pooled standard deviation, with values <0.1 indicating adequate matching.^33^

Odds ratios, comparing odds of prior infection and/or vaccination among cases versus controls, and associated 95% confidence intervals (CIs) were derived using conditional logistic regression, factoring the matching in the study design. This analytical approach minimizes potential bias due to variation in epidemic phase,^20,34^ roll-out of vaccination during the study,^20,34^ or other confounders.^35,36^ CIs were not adjusted for multiplicity and thus should not be used to infer definitive differences between different groups. Interactions were not investigated. Effectiveness measures and associated 95% CIs were calculated by applying the following equation:^20,21,23^

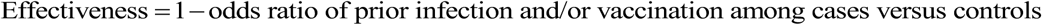

The reference group for all effectiveness estimates comprised individuals with no prior infection and no vaccination. Statistical analyses were conducted in STATA/SE version 17.0 (Stata Corporation, College Station, TX, USA).

### Oversight

Hamad Medical Corporation and Weill Cornell Medicine-Qatar Institutional Review Boards approved this retrospective study with a waiver of informed consent. The study was reported following the Strengthening the Reporting of Observational studies in Epidemiology (STROBE) guidelines. The STROBE checklist can be found in Table S1.

## Results

### Study population

Between December 23, 2021 and February 21, 2022, 1,306,862 individuals received at least two BNT162b2 doses, and 341,438 of these received a booster dose. The median date was May 3, 2021 for the first dose, May 24, 2021 for the second dose, and December 25, 2021 for the booster dose. The median duration between the first and second doses was 21 days (IQR, 21-22 days), and between the second and booster doses was 251 days (IQR, 233-274 days). During the same duration, 893,671 individuals received at least two mRNA-1273 doses, and 135,050 of these received a booster dose. The median date was May 28, 2021 for the first dose, June 27, 2021 for the second dose, and January 12, 2022 for the booster dose. The median duration between the first and second doses was 28 days (IQR, 28-30 days), and between the second and booster doses was 236 days (IQR, 213-260 days).

### Effectiveness against BA.1—the BNT162b2-vaccine study

Figure 1 shows the process for selecting the study populations and Table 1 describes their characteristics. The study was based on the total population of Qatar; thus, the study population is broadly representative of the internationally diverse, but young and predominantly male, total population of Qatar (Table S2).

**Figure 1.**
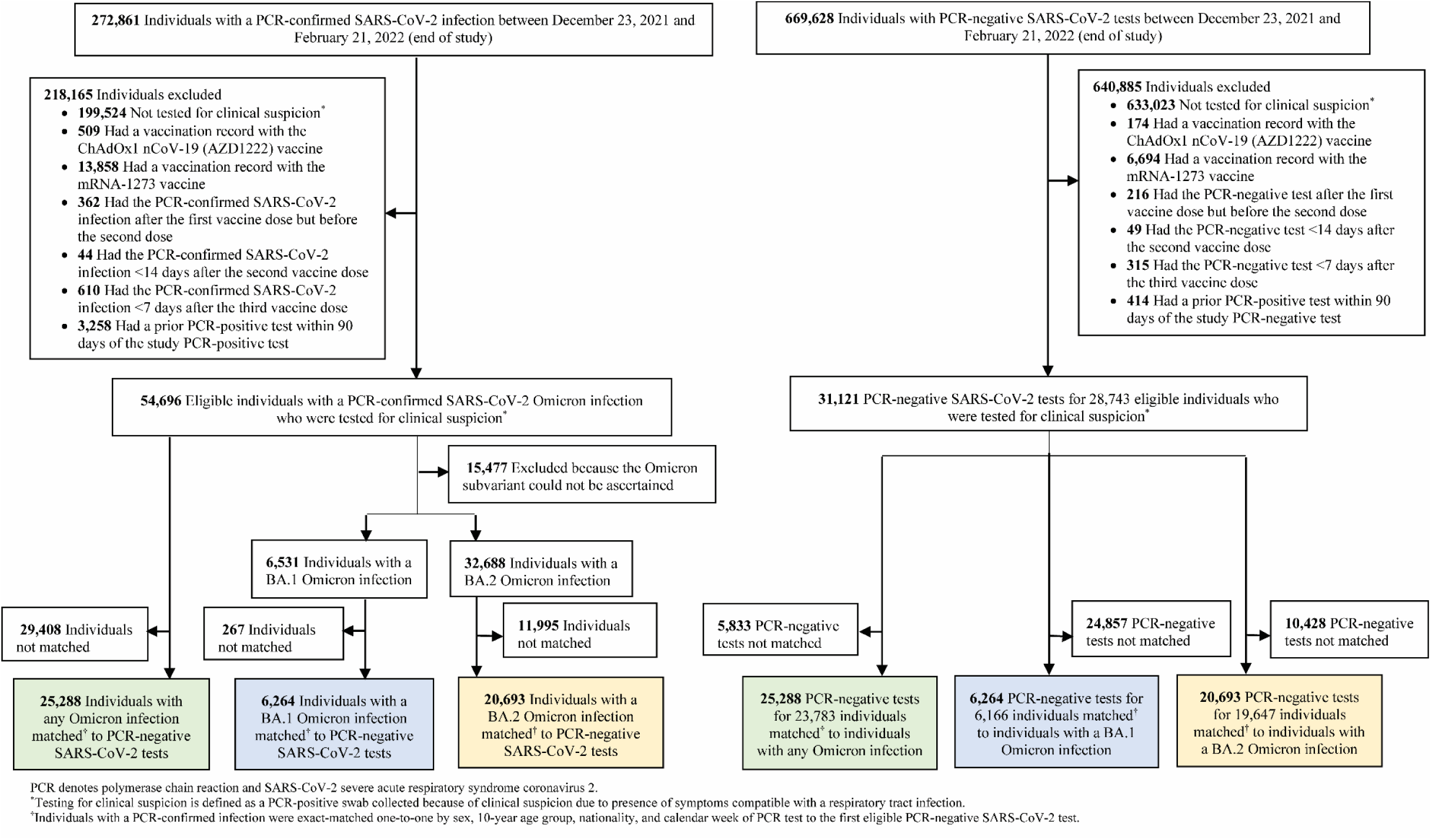
Flowchart describing the population selection process for investigating effectiveness of prior infection, vaccination, and hybrid immunity against symptomatic BA.1 Omicron infection, symptomatic BA.2 Omicron infection, or any Omicron infection in the BNT162b2-vaccine study.

**Table 1.**
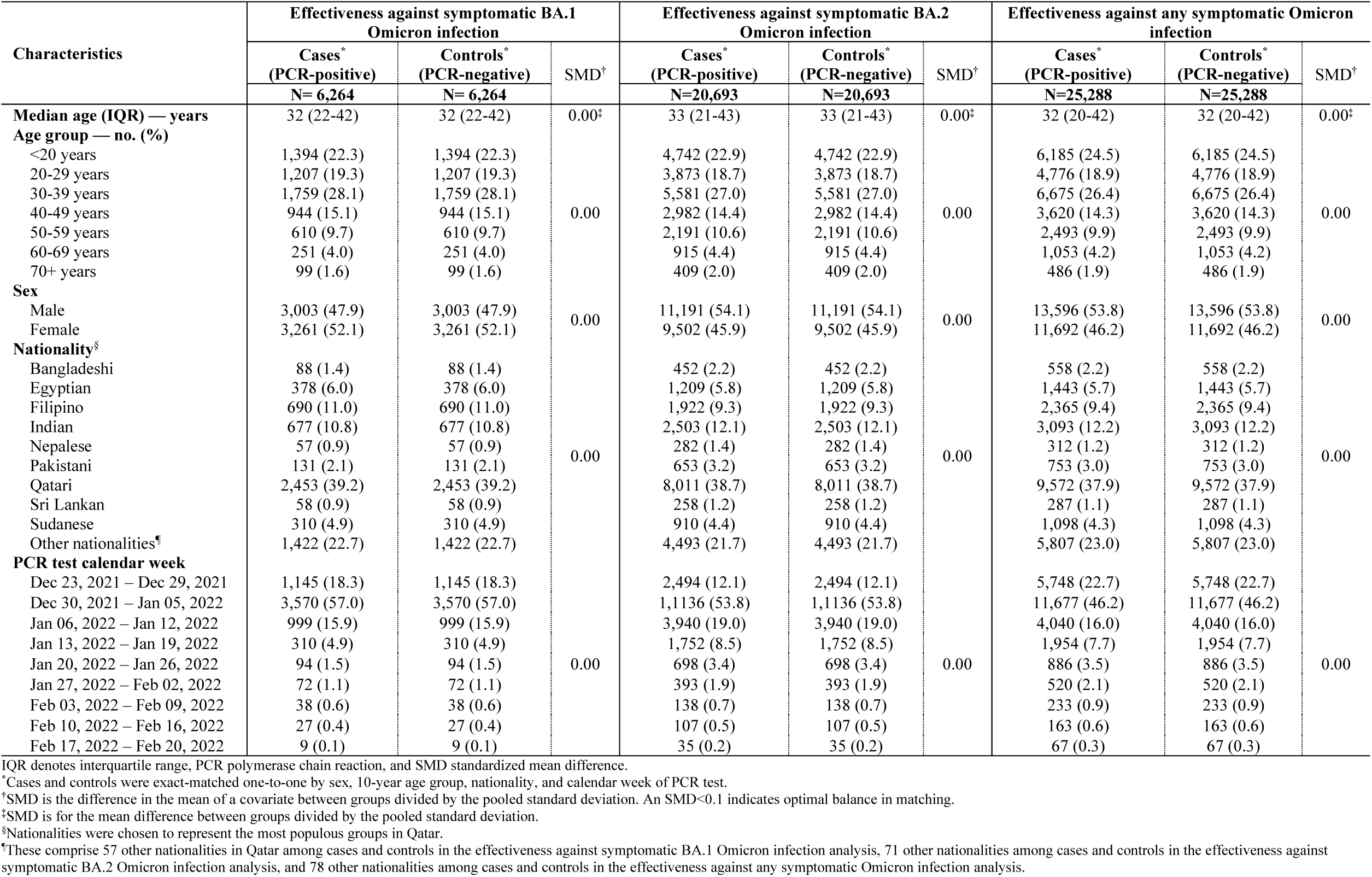
Characteristics of matched cases and controls in samples used to estimate effectiveness against symptomatic BA.1 Omicron infection, symptomatic BA.2 Omicron infection, or any symptomatic Omicron infection in the BNT162b2-vaccine study.

Effectiveness of only prior infection against symptomatic BA.1 infection was 50.2% (95% CI: 38.1-59.9%) (Figure 2A and Table 2). The median duration between the prior infection and the PCR test was 324.5 days (IQR, 274-497 days).

**Figure 2.**
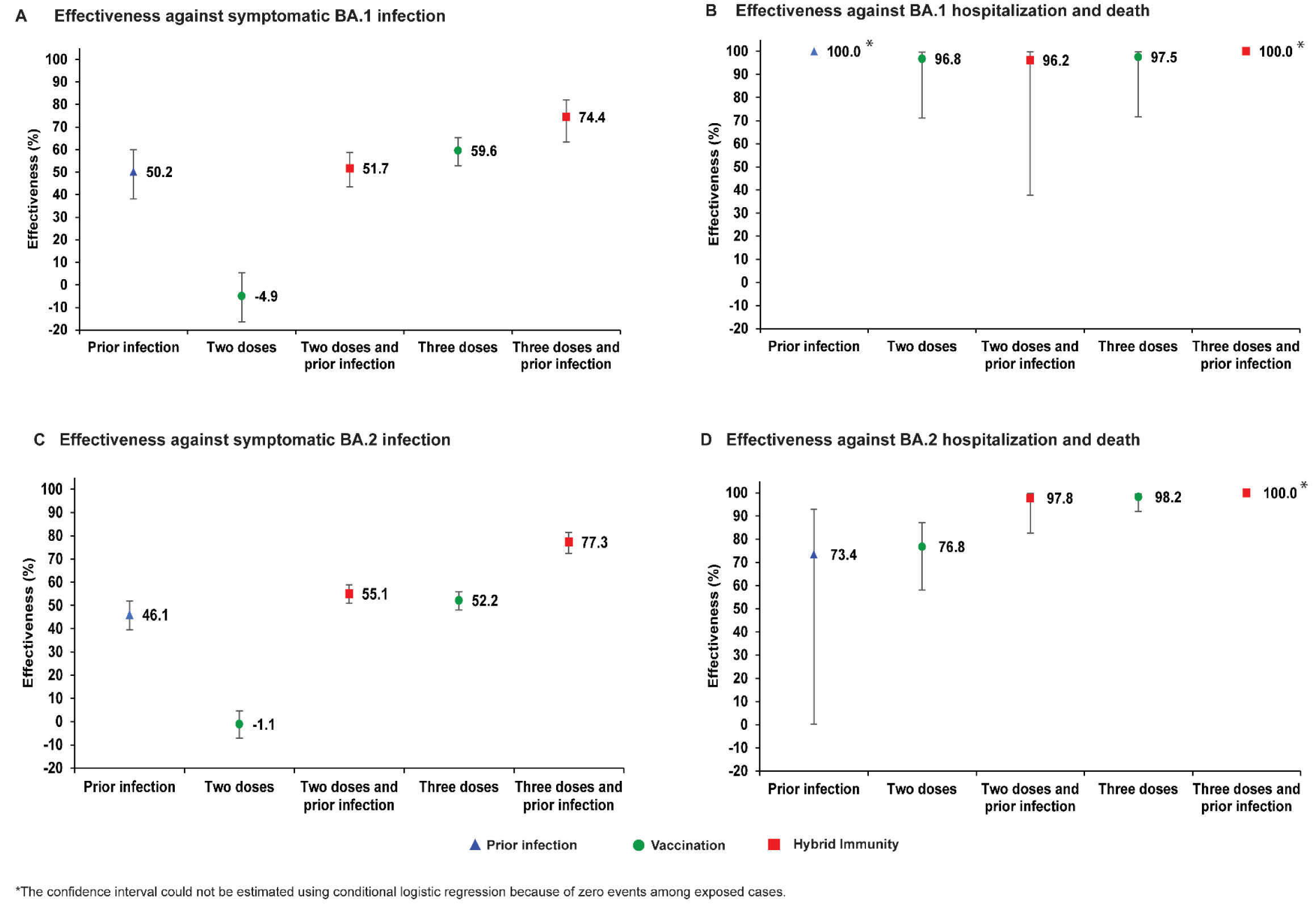
Effectiveness of prior infection, vaccination, and hybrid immunity against symptomatic Omicron infection and against severe, critical, or fatal COVID-19 for the BA.1 (panels A and B, respectively) and BA.2 (panels C and D, respectively) subvariants in the BNT162b2-vaccine study.

**Table 2.** Effectiveness of prior infection, vaccination, and hybrid immunity, in the BNT162b2-vaccine study, against symptomatic infection and against severe, critical, or fatal COVID-19 for BA.1, BA.2, or any Omicron infection.

| Studies | Cases*<br>(PCR-positive) |  | Controls*<br>(PCR-negative) |  | Effectiveness‡ against<br>symptomatic§ infection<br>(95% CI) | Cases†<br>(Severe, critical, or<br>fatal disease)** |  | Controls†<br>(PCR-negative) |  | Effectiveness‡ against<br>hospitalization and death<br>(95% CI) |
| --- | --- | --- | --- | --- | --- | --- | --- | --- | --- | --- |
|  | Exposed† | Unexposed | Exposed† | Unexposed |  | Exposed† | Unexposed | Exposed† | Unexposed |  |
| BA.1 Omicron infection |  |  |  |  |  |  |  |  |  |  |
| Prior infection and no<br>vaccination | 149 | 1,738 | 255 | 1,536 | 50.2<br>(38.1 to 59.9) | 0 | 12 | 6 | 11 | 100.0<br>(Omitted)†† |
| Two doses and no<br>prior infection | 3,449 | 1,738 | 2,762 | 1,536 | -4.9<br>(-16.4 to 5.4) | 5 | 12 | 39 | 11 | 96.8<br>(71.1 to 99.6) |
| Two doses and prior<br>infection | 402 | 1,738 | 688 | 1,536 | 51.7<br>(43.5 to 58.7) | 1 | 12 | 8 | 11 | 96.2<br>(37.7 to 99.8) |
| Three doses and no<br>prior infection | 479 | 1,738 | 892 | 1,536 | 59.6<br>(52.9 to 65.3) | 2 | 12 | 20 | 11 | 97.5<br>(71.7 to 99.8) |
| Three doses and prior<br>infection | 47 | 1,738 | 131 | 1,536 | 74.4<br>(63.4 to 82.2) | 0 | 12 | 7 | 11 | 100.0<br>(Omitted)†† |
| BA.2 Omicron infection |  |  |  |  |  |  |  |  |  |  |
| Prior infection and no<br>vaccination | 565 | 6,051 | 895 | 5,372 | 46.1<br>(39.5 to 51.9) | 3 | 43 | 17 | 50 | 73.4<br>(0.2 to 92.9) |
| Two doses and no<br>prior infection | 10,880 | 6,051 | 8,846 | 5,372 | -1.1<br>(-7.1 to 4.6) | 41 | 43 | 168 | 50 | 76.8<br>(58.0 to 87.1) |
| Two doses and prior<br>infection | 1,160 | 6,051 | 2,108 | 5,372 | 55.1<br>(50.9 to 58.9) | 1 | 43 | 41 | 50 | 97.8<br>(82.6 to 99.7) |
| Three doses and no<br>prior infection | 1,884 | 6,051 | 2,983 | 5,372 | 52.2<br>(48.1 to 55.9) | 3 | 43 | 98 | 50 | 98.2<br>(91.9 to 99.6) |
| Three doses and prior<br>infection | 153 | 6,051 | 489 | 5,372 | 77.3<br>(72.4 to 81.4) | 0 | 43 | 23 | 50 | 100.0<br>(Omitted)†† |
| Any Omicron infection |  |  |  |  |  |  |  |  |  |  |
| Prior infection and no<br>vaccination | 637 | 7,837 | 1,113 | 6,904 | 50.8<br>(45.4 to 55.7) | 4 | 100 | 24 | 139 | 71.6<br>(15.7 to 90.4) |
| Two doses and no<br>prior infection | 13,033 | 7,837 | 10,600 | 6,904 | -0.2<br>(-5.5 to 4.9) | 63 | 100 | 320 | 139 | 73.5<br>(60.5 to 82.2) |
| Two doses and prior<br>infection | 1,360 | 7,837 | 2,501 | 6,904 | 55.5<br>(51.8 to 59.0) | 3 | 100 | 79 | 139 | 94.3<br>(81.3 to 98.3) |
| Three doses and no<br>prior infection | 2,234 | 7,837 | 3,586 | 6,904 | 54.0<br>(50.4 to 57.3) | 12 | 100 | 164 | 139 | 92.5<br>(84.4 to 96.3) |
| Three doses and prior<br>infection | 187 | 7,837 | 584 | 6,904 | 76.3<br>(71.7 to 80.1) | 0 | 100 | 47 | 139 | 100.0<br>(Omitted)†† |

CI denotes confidence interval, COVID-19 coronavirus disease 2019, and PCR polymerase chain reaction.
\*Cases and controls in studies of effectiveness against symptomatic BA.1 Omicron infection, symptomatic BA.2 Omicron infection, and any symptomatic Omicron infection were exact-matched one-to-one by sex, 10-year age group, nationality, and calendar week of PCR test.
<sup>†</sup>Exposed is defined based on outcome: prior infection with no vaccination, vaccination with only two doses but with no prior infection, vaccination with only two doses but with prior infection, vaccination with three doses but with no prior infection, or vaccination with three doses but with prior infection.
<sup>‡</sup>Effectiveness was estimated using the test-negative, case-control study design.<sup>20,21</sup>
<sup>§</sup>A symptomatic infection was defined as a PCR-positive nasopharyngeal swab conducted because of clinical suspicion due to presence of symptoms compatible with a respiratory tract infection.
<sup>†</sup>Cases and controls in studies of effectiveness against COVID-19 hospitalization and death were exact-matched one-to-five by sex, 10-year age group, nationality, and calendar week of PCR test.
<sup>\*\*</sup>Severity,<sup>10</sup> criticality,<sup>10</sup> and fatality<sup>11</sup> were defined according to the World Health Organization guidelines.
\*\*Confidence intervals could not be estimated using conditional logistic regression because of zero events in exposed cases.

Effectiveness of only two-dose (primary series) BNT162b2 vaccination was negligible at -4.9% (95% CI: -16.4-5.4). The median duration between the second dose and the PCR test was 268 days (IQR, 211-293 days), well beyond the duration of protection of two-dose BNT162b2 vaccination against symptomatic Omicron infections.^5,37^ Effectiveness of only three-dose (booster) vaccination was 59.6% (95% CI: 52.9-65.3%). The median duration between the third dose and the PCR test was 42 days (IQR, 27-62 days).

Effectiveness of hybrid immunity of prior infection and two-dose vaccination was 51.7% (95% CI: 43.5-58.7%), similar to effectiveness of only prior infection (Figure 2A and Table 2). Effectiveness of hybrid immunity of prior infection and three-dose vaccination was highest at 74.4% (95% CI: 63.4-82.2%). Meanwhile, prior infection, vaccination, and hybrid immunity all showed strong effectiveness >90% against severe, critical, or fatal COVID-19 due to BA.1 infection, but some of the 95% CIs were wide or could not be estimated (Figure 2B and Table 2).

### Effectiveness against BA.2—the BNT162b2-vaccine study

Effectiveness of only prior infection against symptomatic BA.2 infection was 46.1% (95% CI: 39.5-51.9%) (Figure 2C and Table 2). The median duration between the prior infection and the PCR test was 319 days (IQR, 275-499 days).

Effectiveness of only two-dose BNT162b2 vaccination was negligible at -1.1% (95% CI: -7.1-4.6). The median duration between the second dose and the PCR test was 270 days (IQR, 213-296 days), well beyond the duration of protection of two-dose BNT162b2 vaccination against symptomatic Omicron infections.^5,37^ Effectiveness of only three-dose vaccination was 52.2% (95% CI: 48.1-55.9%). The median duration between the third dose and the PCR test was 43 days (IQR, 26-65 days).

Effectiveness of hybrid immunity of prior infection and two-dose vaccination was 55.1% (95% CI: 50.9-58.9%), similar to effectiveness of only prior infection (Figure 2C and Table 2). Effectiveness of hybrid immunity of prior infection and three-dose vaccination was highest at 77.3% (95% CI: 72.4-81.4%). Meanwhile, prior infection, vaccination, and hybrid immunity all showed strong effectiveness >70% against severe, critical, or fatal COVID-19 due to BA.2 infection, but some of the 95% CIs were wide or could not be estimated (Figure 2D and Table 2).

### Effectiveness against any Omicron infection—the BNT162b2-vaccine study

Effectiveness of prior infection, BNT162b2 vaccination, and hybrid immunity against any symptomatic Omicron infection with either subvariant showed similar levels and patterns to those of effectiveness against BA.1 or BA.2 (Figure 3A and Table 2). Effectiveness of prior infection, vaccination, and hybrid immunity against severe, critical, or fatal COVID-19 due to any Omicron infection also showed similar levels and patterns to those of effectiveness against BA.1 and BA.2 (Figure 3B and Table 2).

**Figure 3.**
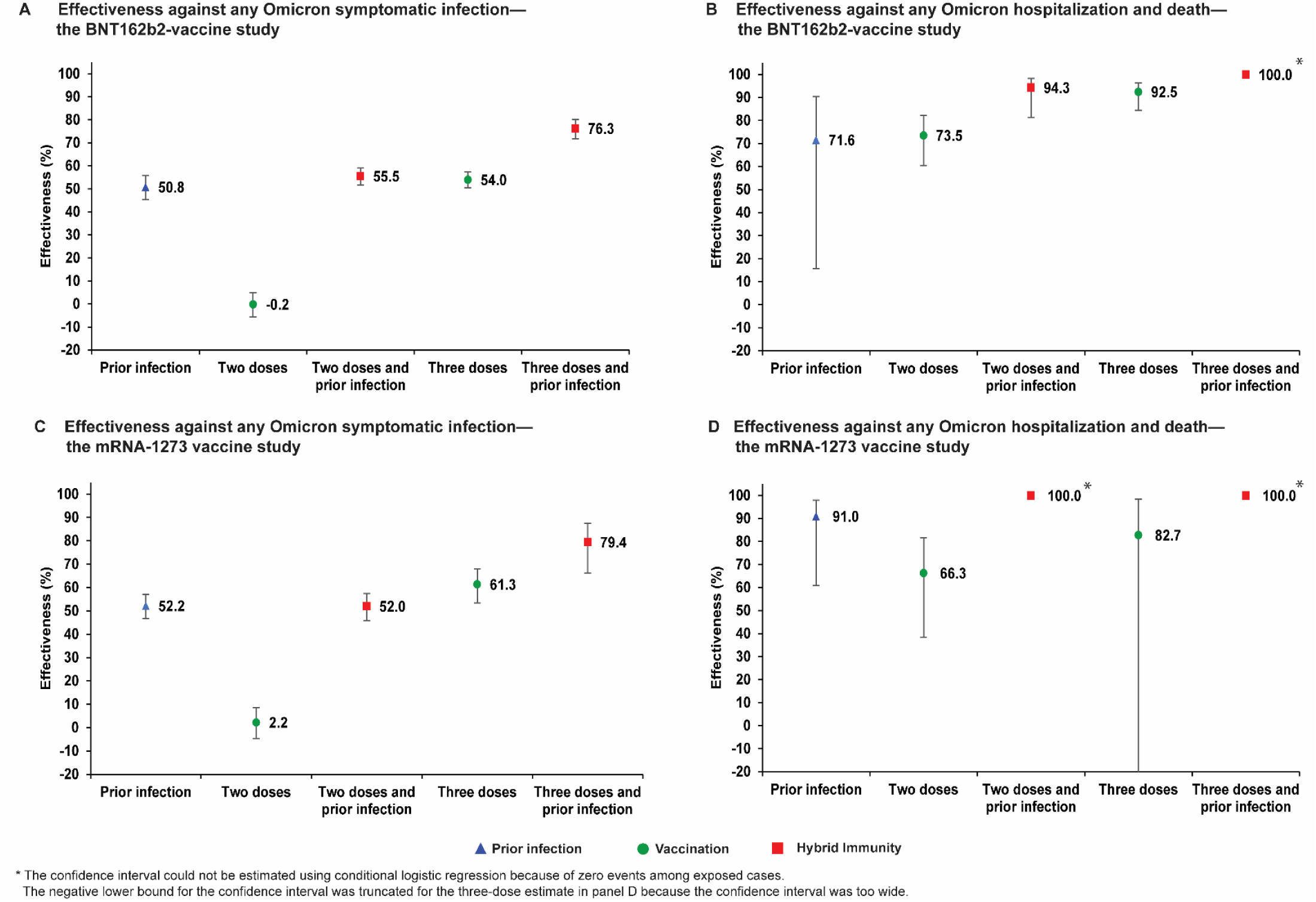
Effectiveness of prior infection, vaccination, and hybrid immunity against any symptomatic Omicron infection and against severe, critical, or fatal COVID-19 in the BNT162b2-vaccine study (panels A and B) and the mRNA-1273-vaccine study (panels C and D).

### Effectiveness against BA.1, BA.2, and any Omicron infection—the mRNA-1273-vaccine study

Figure S2 shows the process for selecting the study populations and Table S3 describes their characteristics. The study population is broadly representative of the population of Qatar (Table S2).

Effectiveness of prior infection, vaccination, and hybrid immunity in the mRNA-1273-vaccine study showed similar levels and patterns to those in the BNT162b2-vaccine study. This was true for effectiveness against BA.1, BA.2, and any Omicron infection, and against both symptomatic infection and severe, critical, or fatal COVID-19 (Figure 4 for BA.1 and BA.2, Figure 3 for any Omicron, and Table S4).

**Figure 4.**
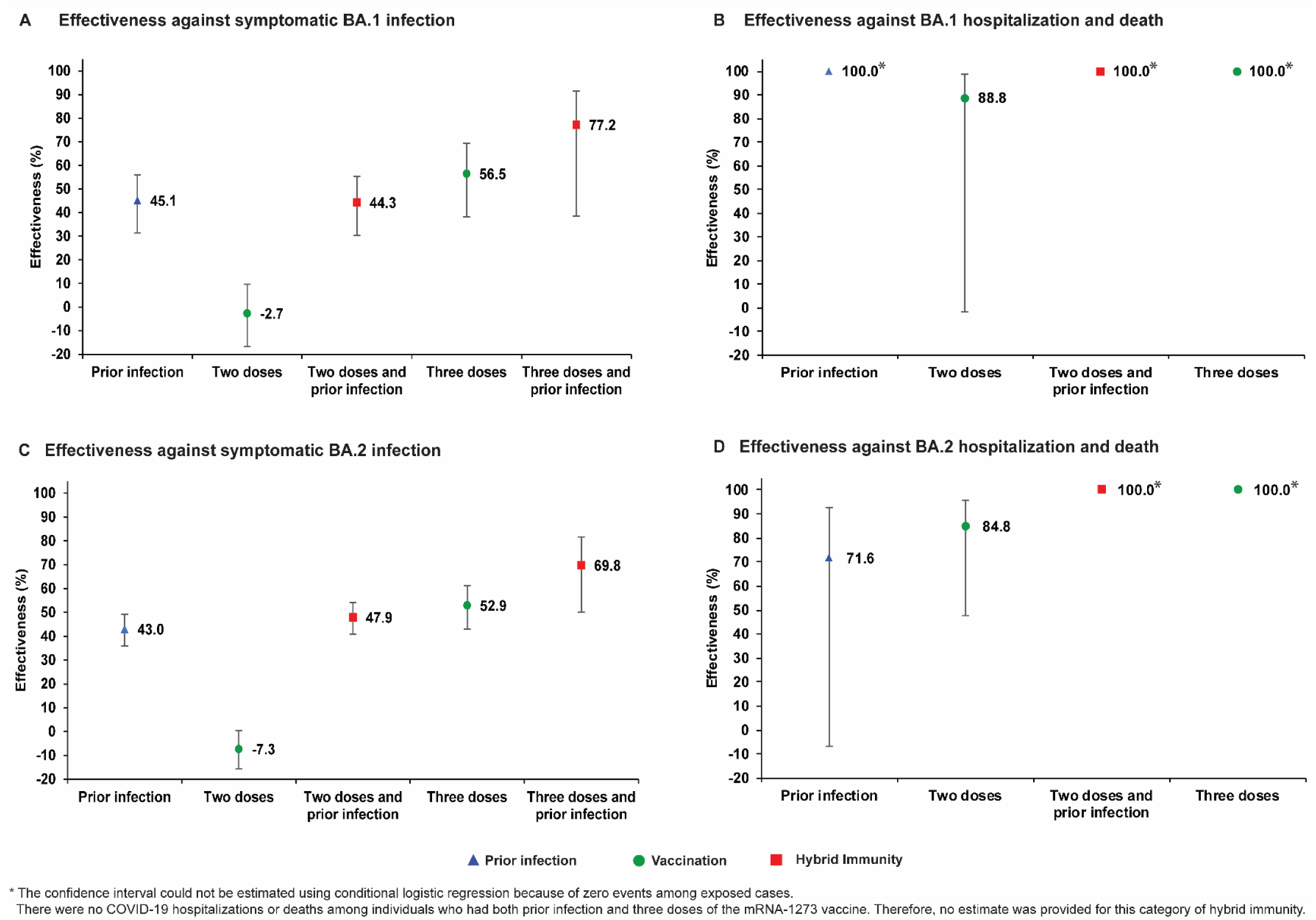
Effectiveness of prior infection, vaccination, and hybrid immunity against symptomatic Omicron infection and against severe, critical, or fatal COVID-19 for the BA.1 (panels A and B, respectively) and BA.2 (panels C and D, respectively) subvariants in the mRNA-1273-vaccine study.

## Discussion

Prior infection with a pre-Omicron variant, nearly a year earlier, was associated with ∼50% reduced risk of infection. There was no discernable difference in protection of prior infection against either BA.1 or BA.2. Meanwhile, two-dose vaccination had negligible effectiveness against either BA.1 or BA.2, but most individuals had received their second doses >8 months earlier. These findings are explained by the short-lived protection of primary-series vaccination against Omicron infections,^5,37^ but more durable protection for natural infection.^3,38^

Booster vaccination was associated with ∼60% reduced risk of infection. There was no discernable difference in protection of booster vaccination against either BA.1 or BA.2. However, most individuals had received their booster doses <45 days earlier, perhaps explaining the relatively high effectiveness.^5^

Protection of hybrid immunity of prior infection and primary-series vaccination was similar to that of prior infection alone, at ∼50%, suggesting that this protection originated from the prior infection and not from primary-series vaccination. This finding is also explained by the short-lived protection of primary-series vaccination against Omicron infections.^5,37^

However, the highest protection was that of hybrid immunity of prior infection and recent booster vaccination, at ∼80%. This finding provides evidence for the benefit of vaccination, even for those with a prior infection. Strikingly, this protection is what one would expect if each of the prior-infection immunity and booster-vaccine immunity acted independently. Since prior infection reduced risk of infection by 50% and booster vaccination reduced it by 60%, the reduction in risk of infection for both combined, if they acted fully independently, is 1− (1− 0.5)×(1− 0.6) = 0.8, that is an 80% reduction, just as observed in this study. This finding suggests that the combined effect of these two forms of immunity reflects neither synergy nor redundancy of their individual biological effects.

Even though these five forms of immunity showed large differences in protection against symptomatic infection that ranged between 0-80%, remarkably, they all showed strong protection against COVID-19 hospitalization and death, at an effectiveness >70% (Figures 2-4). This suggests that any form of prior immunity, whether induced by prior infection or vaccination, is associated with strong and durable protection against COVID-19 hospitalization and death.

There were no discernable differences in the effects of BNT162b2 versus mRNA-1273 vaccination. The results confirmed other findings that we reported recently including a protection of ∼50% for prior infection against reinfection with BA.1,^3^ a protection of ∼50% for mRNA boosters relative to primary series,^6^ and that mRNA vaccines have negligible effectiveness against Omicron several months after the second dose.^5,39^

This study has limitations. BA.1 and BA.2 ascertainment was based on proxy criteria, i.e., presence or absence of an S-gene “target failure” using the TaqPath PCR assay, but this method of ascertainment is well established not only for Omicron subvariants, but also for other variants such as Alpha.^19,40,41^ Some Omicron infections may have been misclassified Delta infections, but this is not likely, as Delta incidence was limited during the study duration (Section S2). While matching was done for sex, age, and nationality, this was not possible for other factors, such as comorbidities, as such data are not available. However, matching by these factors provided demonstrable control of bias in studies of different epidemiologic designs and that used control groups in Qatar.^13,14,18,28,29^ Effectiveness was assessed using an observational, test-negative, case-control, study design, rather than a design in which cohorts of individuals were followed up. However, the cohort study design applied earlier to the same population yielded findings similar to those of the test-negative case-control design,^17,18,42^ supporting the validity of this standard approach in assessing immunity protection.^3,18,20-23^ Moreover, our recent study of effectiveness of boosters relative to primary series used a cohort study design and generated results consistent with the above results.^6^

Nonetheless, one cannot exclude the possibility that in real-world data, bias could arise in unexpected ways, or from unknown sources, such as subtle differences in test-seeking behavior or changes in the pattern of testing with introduction of other testing modalities, such as rapid antigen testing (RAT). For example, with the large Omicron wave in Qatar, use of RAT was expanded to supplement PCR testing starting on January 5, 2022. However, RAT was broadly implemented and probably did not differentially affect PCR testing to introduce bias. With the small proportion of Qatar’s population ≥50 years of age,^12^ our findings may not be generalizable to other countries in which elderly citizens constitute a larger proportion of the population.

Notwithstanding these limitations, findings were consistent with those of other studies for effectiveness against Omicron infection (with no BA.1/BA.2 subvariant specified).^37,43-47^ Moreover, with the mass scale of PCR testing in Qatar,^14^ the likelihood of bias is perhaps minimized. Extensive sensitivity and additional analyses were conducted to investigate effects of potential bias in our earlier studies that used similar methodology. These included different adjustments and controls in the analysis and different study inclusion and exclusion criteria, to investigate whether effectiveness estimates could have been biased.^14,29^ These analyses showed consistent findings.^3,5,14,23,29^

In conclusion, there are no discernable differences in effects of prior infection, vaccination, and hybrid immunity against BA.1 and BA.2. Protection from prior infection with pre-Omicron variants is moderate and durable, but protection of primary-series vaccination is negligible several months after the second dose. Recent booster vaccination has moderate effectiveness, while hybrid immunity from prior infection and recent booster vaccination conferred the strongest protection at ∼80%. All five forms of immunity were associated with strong and durable protection against COVID-19 hospitalization and death.

## Data Availability

The dataset of this study is a property of the Qatar Ministry of Public Health that was provided to the researchers through a restricted-access agreement that prevents sharing the dataset with a third party or publicly. Future access to this dataset can be considered through a direct application for data access to Her Excellency the Minister of Public Health (https://www.moph.gov.qa/english/Pages/default.aspx). Aggregate data are available within the manuscript and its Supplementary information.

## Sources of support and acknowledgements

We acknowledge the many dedicated individuals at Hamad Medical Corporation, the Ministry of Public Health, the Primary Health Care Corporation, Qatar Biobank, Sidra Medicine, and Weill Cornell Medicine-Qatar for their diligent efforts and contributions to make this study possible. The authors are grateful for institutional salary support from the Biomedical Research Program and the Biostatistics, Epidemiology, and Biomathematics Research Core, both at Weill Cornell Medicine-Qatar, as well as for institutional salary support provided by the Ministry of Public Health, Hamad Medical Corporation, and Sidra Medicine. The authors are also grateful for the Qatar Genome Programme and Qatar University Biomedical Research Center for institutional support for the reagents needed for the viral genome sequencing. The funders of the study had no role in study design, data collection, data analysis, data interpretation, or writing of the article. Statements made herein are solely the responsibility of the authors.

## Author contributions

HNA co-designed the study, performed the statistical analyses, and co-wrote the first draft of the article. HC co-designed the study, co-led the statistical analyses, and co-wrote the first draft of the article. LJA conceived and co-designed the study, co-led the statistical analyses, and co-wrote the first draft of the article. PT and MRH conducted the multiplex, RT-qPCR variant screening and viral genome sequencing. HY, FMB, and HAK conducted viral genome sequencing. PC conducted testing of samples using the TaqPath Kit to ascertain the BA.1 versus BA.2 subvariant status of PCR-positive swabs. All authors contributed to data collection and acquisition, database development, discussion and interpretation of the results, and to the writing of the manuscript. All authors have read and approved the final manuscript.

## Supplementary Appendix

**Figure S1.**
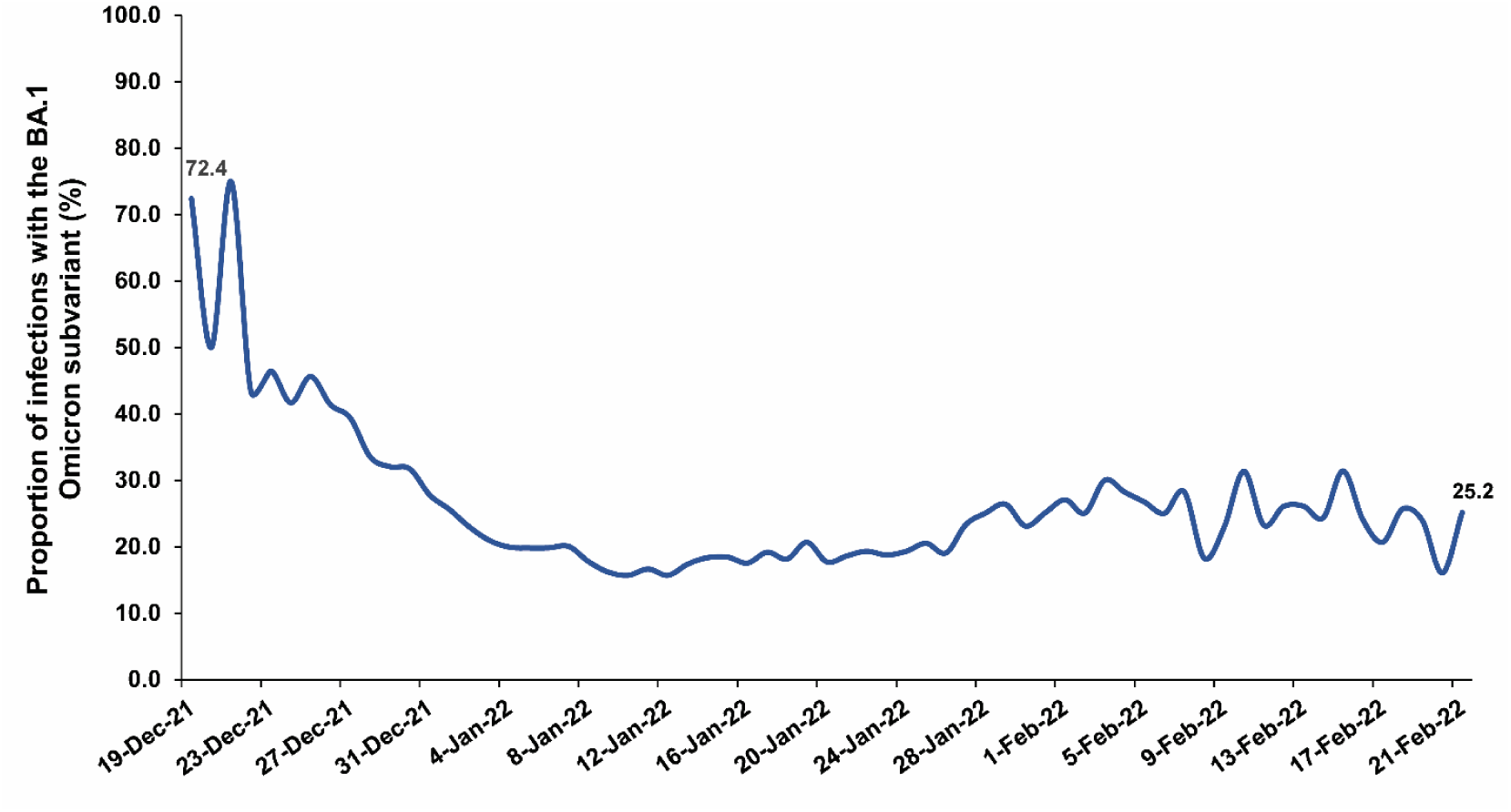
Proportion of Omicron infections with the BA.1 (versus BA.2) subvariant in PCR-positive tests assessed using TaqPath COVID-19 Combo Kit during the study period.

### Table of contents

### Section S1. Coronavirus Disease 2019 (COVID-19) severity, criticality, and fatality classification

Severe Coronavirus Disease 2019 (COVID-19) disease was defined per the World health Organization (WHO) classification as a severe acute respiratory syndrome coronavirus 2 (SARS-CoV-2) infected person with “oxygen saturation of <90% on room air, and/or respiratory rate of >30 breaths/minute in adults and children >5 years old (or ≥60 breaths/minute in children <2 months old or ≥50 breaths/minute in children 2-11 months old or ≥40 breaths/minute in children 1–5 years old), and/or signs of severe respiratory distress (accessory muscle use and inability to complete full sentences, and, in children, very severe chest wall indrawing, grunting, central cyanosis, or presence of any other general danger signs)”.^1^ Detailed WHO criteria for classifying SARS-CoV-2 infection severity can be found in the WHO technical report.^1^

Critical COVID-19 disease was defined per WHO classification as a SARS-CoV-2 infected person with “acute respiratory distress syndrome, sepsis, septic shock, or other conditions that would normally require the provision of life sustaining therapies such as mechanical ventilation (invasive or non-invasive) or vasopressor therapy”.^1^ Detailed WHO criteria for classifying SARS-CoV-2 infection criticality can be found in the WHO technical report.^1^

COVID-19 death was defined per WHO classification as “a death resulting from a clinically compatible illness, in a probable or confirmed COVID-19 case, unless there is a clear alternative cause of death that cannot be related to COVID-19 disease (e.g. trauma). There should be no period of complete recovery from COVID-19 between illness and death. A death due to COVID-19 may not be attributed to another disease (e.g. cancer) and should be counted independently of preexisting conditions that are suspected of triggering a severe course of COVID-19”. Detailed WHO criteria for classifying COVID-19 death can be found in the WHO technical report.^2^

### Section S2. Laboratory methods and subvariant ascertainment

#### Real-time reverse-transcription polymerase chain reaction testing

Nasopharyngeal and/or oropharyngeal swabs were collected for PCR testing and placed in Universal Transport Medium (UTM). Aliquots of UTM were: 1) extracted on KingFisher Flex (Thermo Fisher Scientific, USA), MGISP-960 (MGI, China), or ExiPrep 96 Lite (Bioneer, South Korea) followed by testing with real-time reverse-transcription PCR (RT-qPCR) using TaqPath COVID-19 Combo Kits (Thermo Fisher Scientific, USA) on an ABI 7500 FAST (Thermo Fisher Scientific, USA); 2) tested directly on the Cepheid GeneXpert system using the Xpert Xpress SARS-CoV-2 (Cepheid, USA); or 3) loaded directly into a Roche cobas 6800 system and assayed with the cobas SARS-CoV-2 Test (Roche, Switzerland). The first assay targets the viral S, N, and ORF1ab gene regions. The second targets the viral N and E-gene regions, and the third targets the ORF1ab and E-gene regions.

All PCR testing was conducted at the Hamad Medical Corporation Central Laboratory or Sidra Medicine Laboratory, following standardized protocols.

#### Classification of infections by subvariant type

Surveillance for SARS-CoV-2 variants in Qatar is mainly based on viral genome sequencing and multiplex RT-qPCR variant screening^3^ of random positive clinical samples,^4-9^ complemented by deep sequencing of wastewater samples.^6,10^

A total of 315 random SARS-CoV-2-positive specimens collected between December 19, 2021 and January 22, 2022 were viral whole-genome sequenced on a Nanopore GridION sequencing device. Of these, 300 (95.2%) were confirmed as Omicron infections and 15 (4.8%) as Delta (B.1.617.2)^11^ infections.^6,12-15^ Of 286 Omicron infections with confirmed subvariant status, 68 (23.8%) were BA.1 cases and 218 (76.2%) were BA.2 cases.

Additionally, a total of 1,315 random SARS-CoV-2-positive specimens collected between December 22, 2021 and January 1, 2022 were RT-qPCR genotyped. The RT-qPCR genotyping identified 1 B.1.617.2-like Delta case, 366 BA.1-like Omicron cases, 898 BA.2-like Omicron cases, and 50 were undetermined cases where the genotype could not be assigned.

The accuracy of the RT-qPCR genotyping was verified against either Sanger sequencing of the receptor-binding domain (RBD) of SARS-CoV-2 surface glycoprotein (S) gene, or by viral whole-genome sequencing on a Nanopore GridION sequencing device. From 147 random SARS-CoV-2-positive specimens all collected in December of 2021, RT-qPCR genotyping was able to assign a genotype in 129 samples. The agreement between RT-qPCR genotyping and sequencing was 100% for Delta (n=82), 100% for Omicron BA.1 (n=18), and 93% for Omicron BA.2 (27 of 29 were correctly assigned to BA.2 and remaining 2 specimens genotyped as BA.2 were B.1.617.2 by sequencing). Of the remaining 18 specimens: 10 failed PCR amplification and sequencing, 8 could not be assigned a genotype by RT-qPCR (4 of 8 were B.1.617.2 by sequencing, and the remaining 4 failed sequencing). All the variant RT-qPCR genotyping was conducted at the Sidra Medicine Laboratory following standardized protocols.

The large Omicron-wave exponential-growth phase in Qatar started on December 19, 2021 and peaked in mid-January, 2022.^6,12-15^ The study duration coincided with the intense Omicron wave where Delta incidence was limited. Accordingly, any PCR-positive test during the study duration, between December 23, 2021 and February 21, 2022, was assumed to be an Omicron infection. Of note that the study duration started on December 23, 2021, and not on December 19, 2021, to minimize the occurrence of residual Delta incidence during the first few days of the Omicron wave.

Informed by the viral genome sequencing and the RT-qPCR genotyping, a SARS-CoV-2 infection with the BA.1 subvariant was proxied as an S-gene “target failure” (SGTF) case using the TaqPath COVID-19 Combo Kit (Thermo Fisher Scientific, USA) that tests for the S-gene and is affected by the del69/70 mutation in the S-gene.^16^ A SARS-CoV-2 infection with the BA.2 subvariant was proxied as a non-SGTF case using this TaqPath Kit.

**Table S1.**
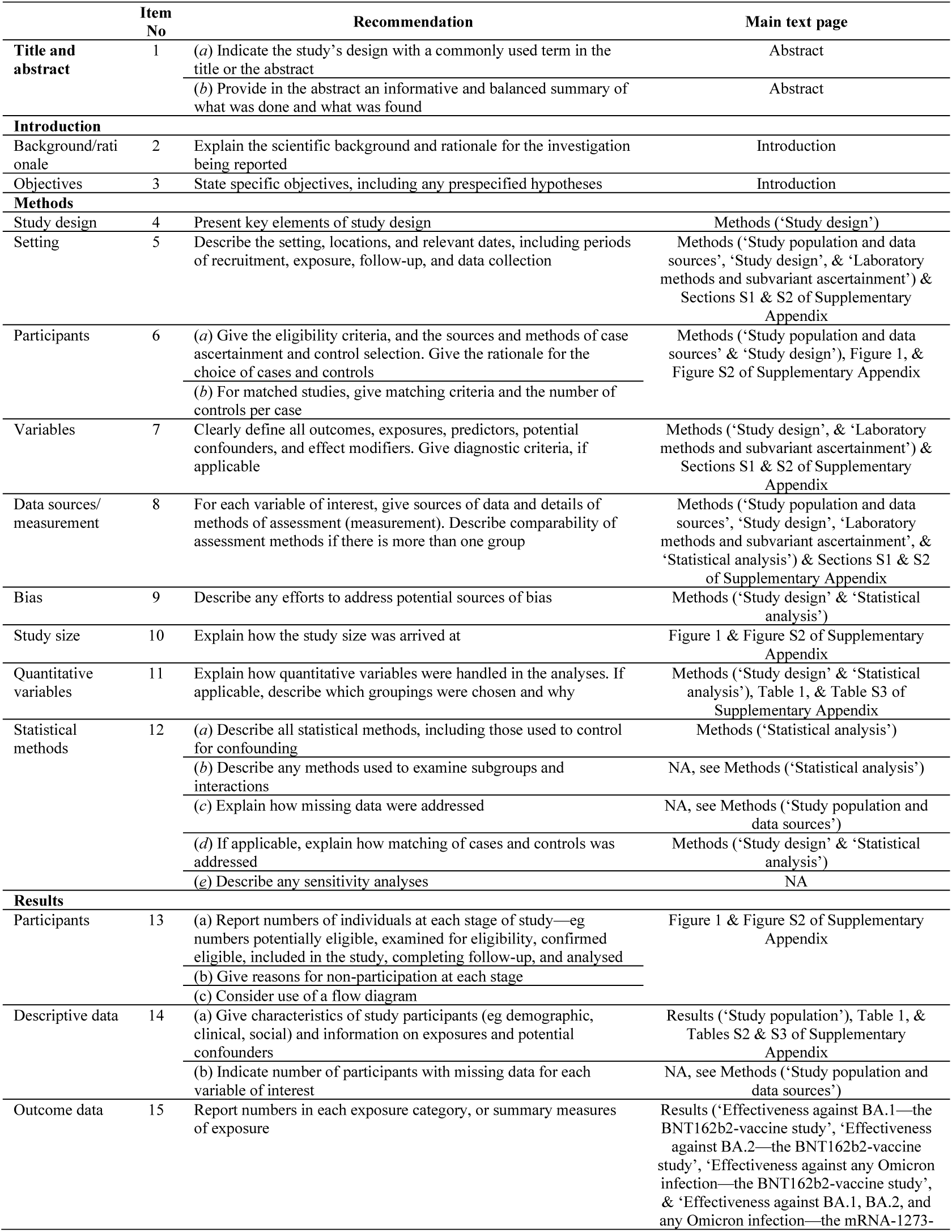
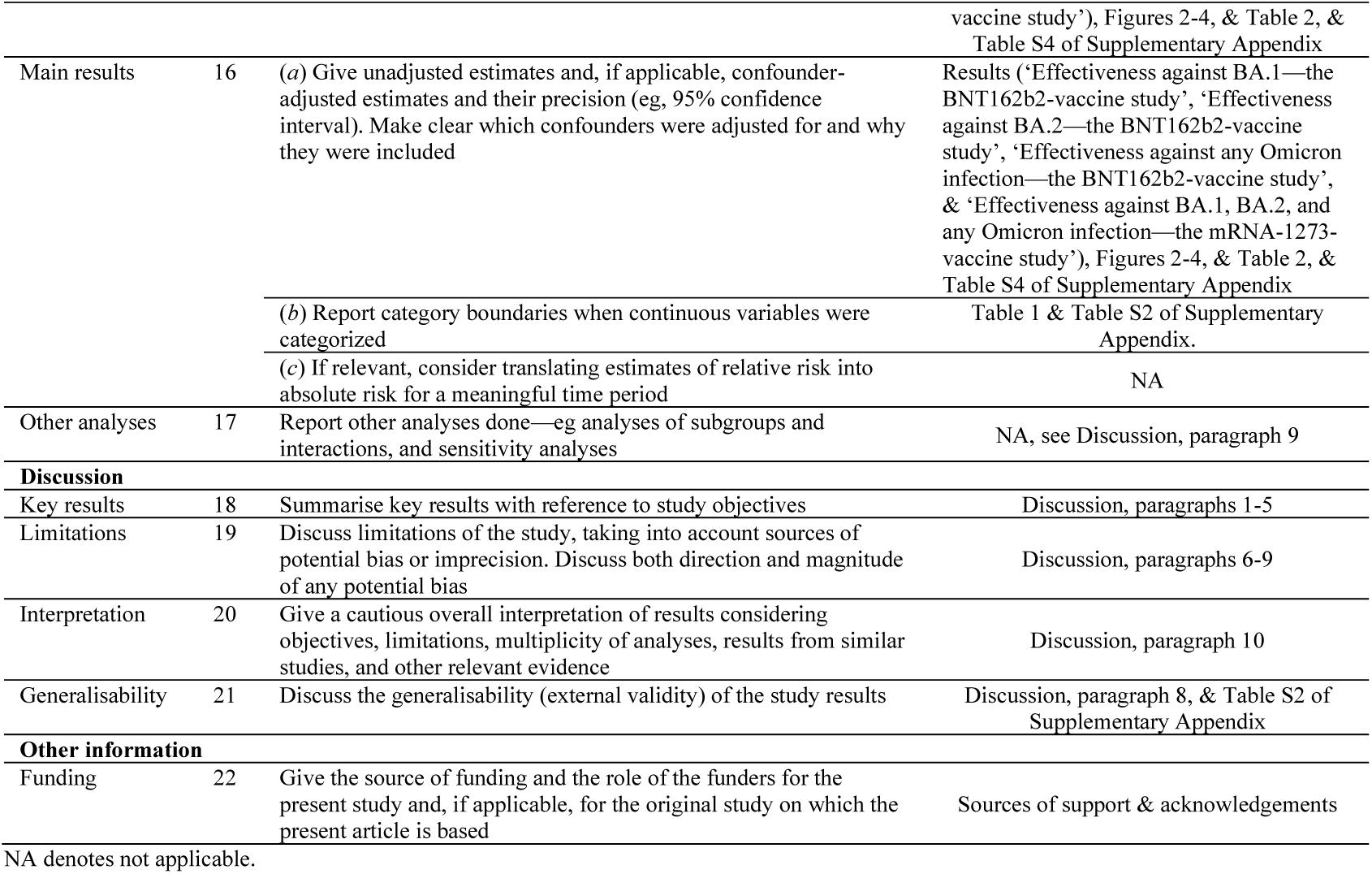
Strengthening the Reporting of Observational studies in Epidemiology (STROBE) checklist for case-control studies.

**Table S2.**
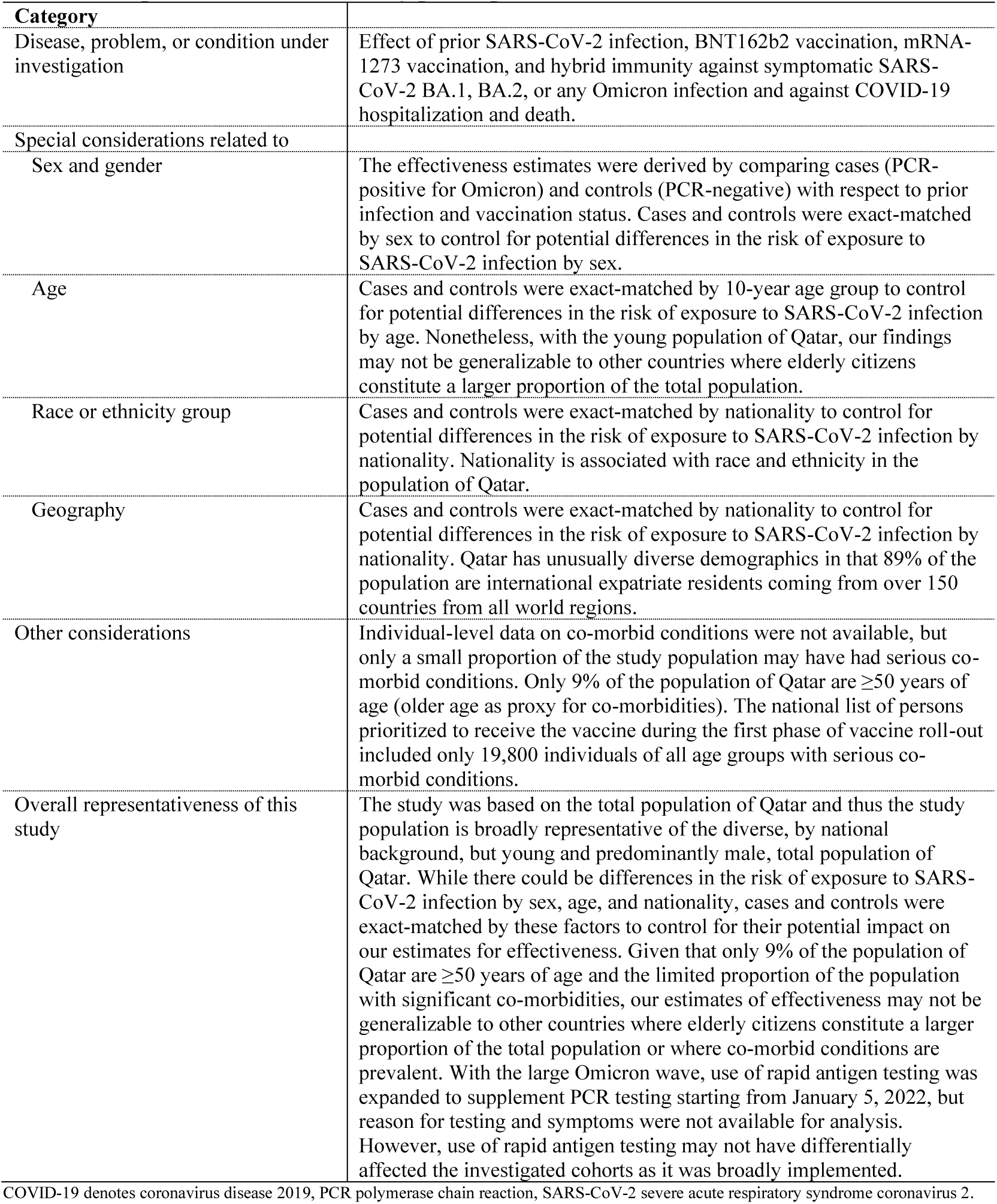
Representativeness of study participants.

| Category |  |
| --- | --- |
| Disease, problem, or condition under investigation | Effect of prior SARS-CoV-2 infection, BNT162b2 vaccination, mRNA-1273 vaccination, and hybrid immunity against symptomatic SARS-CoV-2 BA.1, BA.2, or any Omicron infection and against COVID-19 hospitalization and death. |
| Special considerations related to |  |
| Sex and gender | The effectiveness estimates were derived by comparing cases (PCR-positive for Omicron) and controls (PCR-negative) with respect to prior infection and vaccination status. Cases and controls were exact-matched by sex to control for potential differences in the risk of exposure to SARS-CoV-2 infection by sex. |
| Age | Cases and controls were exact-matched by 10-year age group to control for potential differences in the risk of exposure to SARS-CoV-2 infection by age. Nonetheless, with the young population of Qatar, our findings may not be generalizable to other countries where elderly citizens constitute a larger proportion of the total population. |
| Race or ethnicity group | Cases and controls were exact-matched by nationality to control for potential differences in the risk of exposure to SARS-CoV-2 infection by nationality. Nationality is associated with race and ethnicity in the population of Qatar. |
| Geography | Cases and controls were exact-matched by nationality to control for potential differences in the risk of exposure to SARS-CoV-2 infection by nationality. Qatar has unusually diverse demographics in that 89% of the population are international expatriate residents coming from over 150 countries from all world regions. |
| Other considerations | Individual-level data on co-morbid conditions were not available, but only a small proportion of the study population may have had serious co-morbid conditions. Only 9% of the population of Qatar are $\geq 50$ years of age (older age as proxy for co-morbidities). The national list of persons prioritized to receive the vaccine during the first phase of vaccine roll-out included only 19,800 individuals of all age groups with serious co-morbid conditions. |
| Overall representativeness of this study | The study was based on the total population of Qatar and thus the study population is broadly representative of the diverse, by national background, but young and predominantly male, total population of Qatar. While there could be differences in the risk of exposure to SARS-CoV-2 infection by sex, age, and nationality, cases and controls were exact-matched by these factors to control for their potential impact on our estimates for effectiveness. Given that only 9% of the population of Qatar are $\geq 50$ years of age and the limited proportion of the population with significant co-morbidities, our estimates of effectiveness may not be generalizable to other countries where elderly citizens constitute a larger proportion of the total population or where co-morbid conditions are prevalent. With the large Omicron wave, use of rapid antigen testing was expanded to supplement PCR testing starting from January 5, 2022, but reason for testing and symptoms were not available for analysis. However, use of rapid antigen testing may not have differentially affected the investigated cohorts as it was broadly implemented. |

COVID-19 denotes coronavirus disease 2019, PCR polymerase chain reaction, SARS-CoV-2 severe acute respiratory syndrome coronavirus 2.

**Table S3.** Characteristics of matched cases and controls in samples used to estimate effectiveness against symptomatic BA.1 Omicron infection, symptomatic BA.2 Omicron infection, or any symptomatic Omicron infection in the mRNA-1273-vaccine study.

| Characteristics | Effectiveness against symptomatic BA.1 Omicron infection |  |  | Effectiveness against symptomatic BA.2 Omicron infection |  |  | Effectiveness against any symptomatic Omicron infection |  |  |
| --- | --- | --- | --- | --- | --- | --- | --- | --- | --- |
|  | Cases*<br>(PCR-positive) | Controls*<br>(PCR-negative) | SMD<sup>†</sup> | Cases*<br>(PCR-positive) | Controls*<br>(PCR-negative) | SMD<sup>†</sup> | Cases*<br>(PCR-positive) | Controls*<br>(PCR-negative) | SMD<sup>†</sup> |
|  | N= 3,212 | N= 3,212 |  | N=10,646 | N=10,646 |  | N=13,139 | N=13,139 |  |
| **Median age (IQR) — years** | 30 (15-38) | 30 (14-38) | 0.01<sup>‡</sup> | 29 (11-38) | 29 (11-38) | 0.01<sup>‡</sup> | 28 (9-37) | 29 (9-38) | 0.00<sup>‡</sup> |
| **Age group — no. (%)** |  |  |  |  |  |  |  |  |  |
| <20 years | 874 (27.2) | 874 (27.2) |  | 3,264 (30.7) | 3,264 (30.7) |  | 4,375 (33.3) | 4,375 (33.3) |  |
| 20-29 years | 678 (21.1) | 678 (21.1) |  | 2,132 (20.0) | 2,132 (20.0) |  | 2,556 (19.5) | 2,556 (19.5) |  |
| 30-39 years | 937 (29.2) | 937 (29.2) |  | 2,903 (27.3) | 2,903 (27.3) |  | 3,412 (26.0) | 3,412 (26.0) |  |
| 40-49 years | 435 (13.5) | 435 (13.5) | 0.00 | 1,328 (12.5) | 1,328 (12.5) | 0.00 | 1,609 (12.2) | 1,609 (12.2) | 0.00 |
| 50-59 years | 214 (6.7) | 214 (6.7) |  | 720 (6.8) | 720 (6.8) |  | 820 (6.2) | 820 (6.2) |  |
| 60-69 years | 55 (1.7) | 55 (1.7) |  | 197 (1.9) | 197 (1.9) |  | 235 (1.8) | 235 (1.8) |  |
| 70+ years | 19 (0.6) | 19 (0.6) |  | 102 (1.0) | 102 (1.0) |  | 132 (1.0) | 132 (1.0) |  |
| **Sex** |  |  |  |  |  |  |  |  |  |
| Male | 1,570 (48.9) | 1,570 (48.9) | 0.00 | 6,117 (57.5) | 6,117 (57.5) | 0.00 | 7,386 (56.2) | 7,386 (56.2) | 0.00 |
| Female | 1,642 (51.1) | 1,642 (51.1) |  | 4,529 (42.5) | 4,529 (42.5) |  | 5,753 (43.8) | 5,753 (43.8) |  |
| **Nationality<sup>§</sup>** |  |  |  |  |  |  |  |  |  |
| Bangladeshi | 68 (2.1) | 68 (2.1) |  | 397 (3.7) | 397 (3.7) |  | 493 (3.8) | 493 (3.8) |  |
| Egyptian | 212 (6.6) | 212 (6.6) |  | 606 (5.7) | 606 (5.7) |  | 749 (5.7) | 749 (5.7) |  |
| Filipino | 487 (15.2) | 487 (15.2) |  | 1,183 (11.1) | 1,183 (11.1) |  | 1,409 (10.7) | 1,409 (10.7) |  |
| Indian | 450 (14.0) | 450 (14.0) |  | 1,618 (15.2) | 1,618 (15.2) |  | 2,028 (15.4) | 2,028 (15.4) |  |
| Nepalese | 54 (1.7) | 54 (1.7) | 0.00 | 281 (2.6) | 281 (2.6) | 0.00 | 305 (2.3) | 305 (2.3) | 0.00 |
| Pakistani | 103 (3.2) | 103 (3.2) |  | 509 (4.8) | 509 (4.8) |  | 581 (4.4) | 581 (4.4) |  |
| Qatari | 768 (23.9) | 768 (23.9) |  | 2,702 (25.4) | 2,702 (25.4) |  | 3,319 (25.3) | 3,319 (25.3) |  |
| Sri Lankan | 40 (1.2) | 40 (1.2) |  | 216 (2.0) | 216 (2.0) |  | 238 (1.8) | 238 (1.8) |  |
| Sudanese | 200 (6.2) | 200 (6.2) |  | 646 (6.1) | 646 (6.1) |  | 774 (5.9) | 774 (5.9) |  |
| Other nationalities<sup>¶</sup> | 830 (25.8) | 830 (25.8) |  | 2,488 (23.4) | 2,488 (23.4) |  | 3,243 (24.7) | 3,243 (24.7) |  |
| **PCR test calendar week** |  |  |  |  |  |  |  |  |  |
| Dec 23, 2021 – Dec 29, 2021 | 638 (19.9) | 638 (19.9) |  | 1,464 (13.8) | 1,464 (13.8) |  | 3,173 (24.1) | 3,173 (24.1) |  |
| Dec 30, 2021 – Jan 05, 2022 | 1,865 (58.1) | 1,865 (58.1) |  | 5,800 (54.5) | 5,800 (54.5) |  | 6,085 (46.3) | 6,085 (46.3) |  |
| Jan 06, 2022 – Jan 12, 2022 | 468 (14.6) | 468 (14.6) |  | 1,966 (18.5) | 1,966 (18.5) |  | 2,055 (15.6) | 2,055 (15.6) |  |
| Jan 13, 2022 – Jan 19, 2022 | 130 (4.0) | 130 (4.0) |  | 856 (8.0) | 856 (8.0) |  | 996 (7.6) | 996 (7.6) |  |
| Jan 20, 2022 – Jan 26, 2022 | 45 (1.4) | 45 (1.4) | 0.00 | 275 (2.6) | 275 (2.6) | 0.00 | 384 (2.9) | 384 (2.9) | 0.00 |
| Jan 27, 2022 – Feb 02, 2022 | 34 (1.1) | 34 (1.1) |  | 154 (1.4) | 154 (1.4) |  | 223 (1.7) | 223 (1.7) |  |
| Feb 03, 2022 – Feb 09, 2022 | 19 (0.6) | 19 (0.6) |  | 68 (0.6) | 68 (0.6) |  | 122 (0.9) | 122 (0.9) |  |
| Feb 10, 2022 – Feb 16, 2022 | 12 (0.4) | 12 (0.4) |  | 50 (0.5) | 50 (0.5) |  | 77 (0.6) | 77 (0.6) |  |
| Feb 17, 2022 – Feb 20, 2022 | 1 (0.03) | 1 (0.03) |  | 13 (0.1) | 13 (0.1) |  | 24 (0.2) | 24 (0.2) |  |

IQR denotes interquartile range, PCR polymerase chain reaction and SMD standardized mean difference.
\*Cases and controls were exact-matched one-to-one by sex, 10-year age group, nationality, and calendar week of PCR test.
<sup>†</sup>SMD is the difference in the mean of a covariate between groups divided by the pooled standard deviation. An SMD<0.1 indicates optimal balance in matching.
<sup>‡</sup>SMD is for the mean difference between groups divided by the pooled standard deviation.
<sup>§</sup>Nationalities were chosen to represent the most populous groups in Qatar.
<sup>¶</sup>These comprise 42 other nationalities in Qatar among cases and controls in the effectiveness against symptomatic BA.1 Omicron infection analysis, 63 other nationalities among cases and controls in the effectiveness against symptomatic BA.2 Omicron infection analysis, and 69 other nationalities among cases and controls in the effectiveness against any symptomatic Omicron infection analysis.

**Figure S2.**
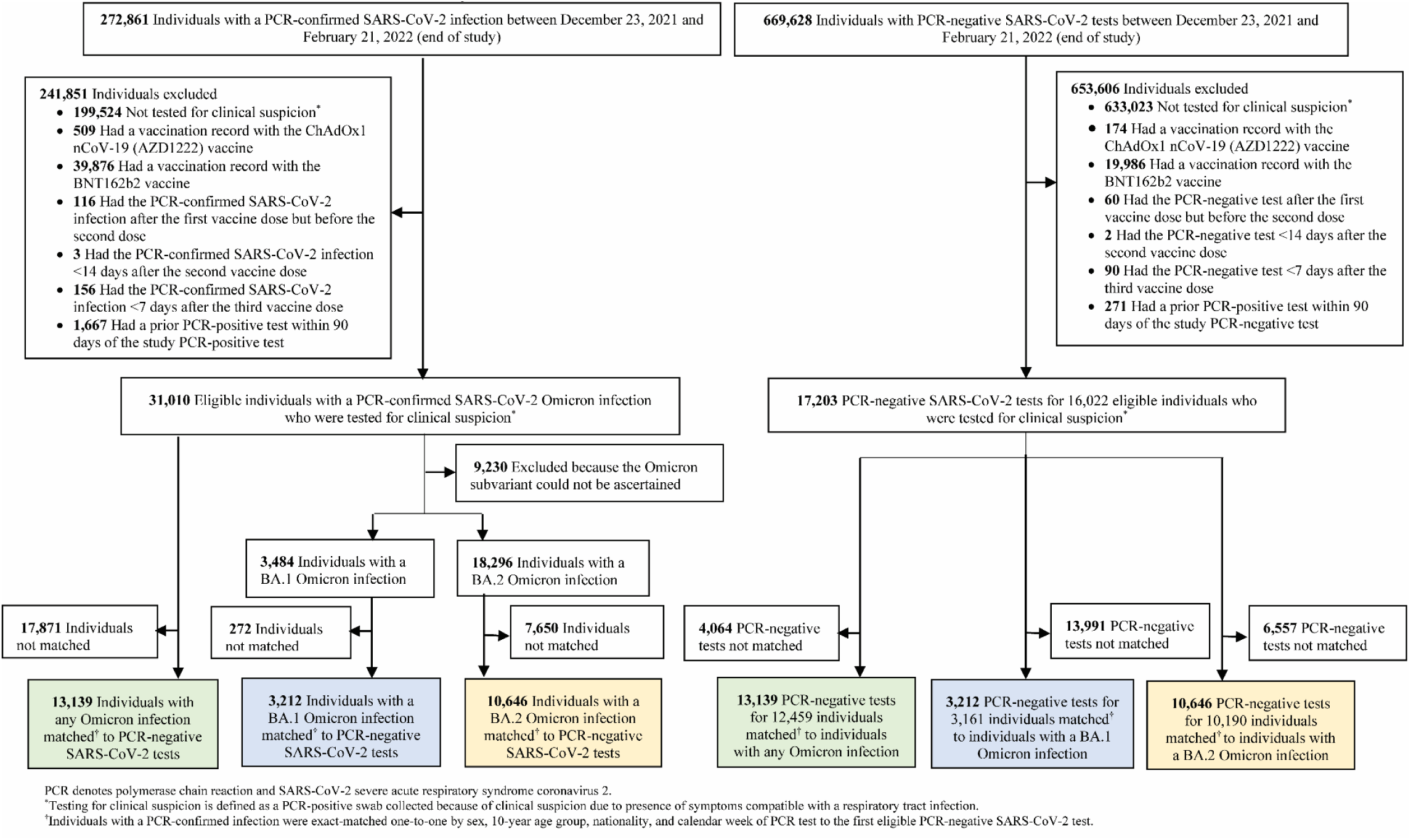
Flowchart describing the population selection process for investigating effectiveness of prior infection, vaccination, and hybrid immunity against symptomatic BA.1 Omicron infection, symptomatic BA.2 Omicron infection, or any Omicron infection in the mRNA-1273-vaccine study.

**Table S4.**
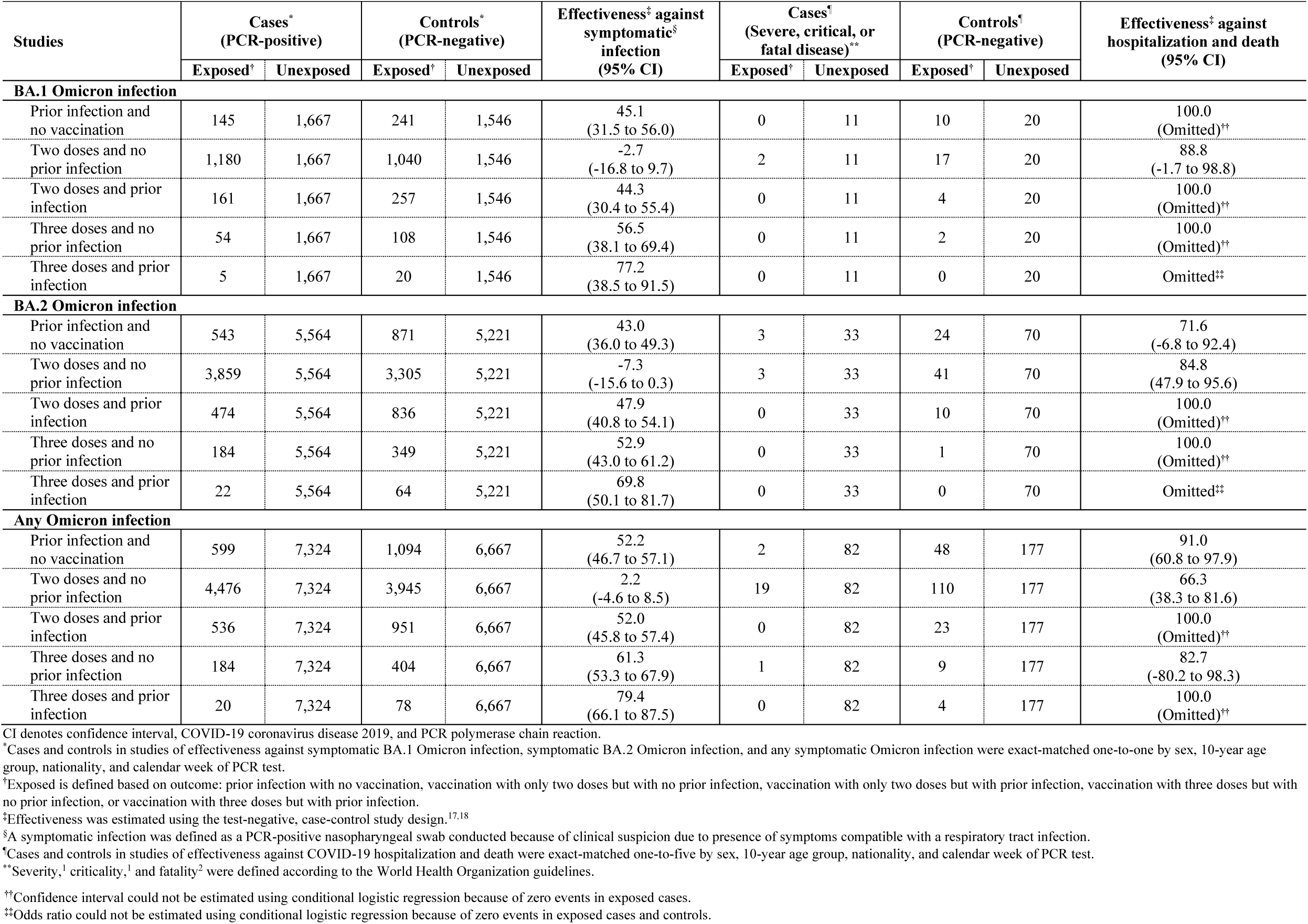
Effectiveness of prior infection, vaccination, and hybrid immunity, in the mRNA-1273-vaccine study, against symptomatic infection and against severe, critical, or fatal COVID-19 for BA.1, BA.2, or any Omicron infection.

